# Detection of SARS-CoV-2 antibodies using commercial assays and seroconversion patterns in hospitalized patients

**DOI:** 10.1101/2020.05.04.20090027

**Authors:** E Tuaillon, K Bolloré, A Pisoni, S Debiesse, C Renault, S Marie, S Groc, C Niels, N Pansu, AM Dupuy, D Morquin, V Foulongne, A Bourdin, V Le Moing, P Van de Perre

**Affiliations:** Pathogenesis and Control of Chronic Infections, University of Montpellier, INSERM, EFS; CHU Montpellier, Montpellier, France; Pathogenesis and Control of Chronic Infections, University of Montpellier, INSERM, EFS, Montpellier, France; Montpellier University Hospital, Montpellier, France

**Keywords:** COVID-19, SARS-CoV-2 antibodies, point of care tests, ELISA

## Abstract

SARS-CoV-2 antibody assays are needed for serological surveys and as a complement to molecular tests to confirm COVID-19. However, the kinetics of the humoral response against SARS-CoV-2 remains poorly described and relies on the performance of the different serological tests.

In this study, we evaluated the performance of six CE-marked point-of-care tests (POC) and three ELISA assays for the diagnosis of COVID-19 by exploring seroconversions in hospitalized patients who tested positive for SARS-CoV-2 RNA.

Both the ELISA and POC tests were able to detect SARS-CoV-2 antibodies in at least half of the samples collected seven days or more after the onset of symptoms. After 15 days, the rate of detection rose to over 80% but without reaching 100%, irrespective of the test used. More than 90% of the samples collected after 15 days tested positive using the iSIA and Accu-Tell^®^ POC tests and the ID.Vet IgG ELISA assay. Seroconversion was observed 5 to 12 days after the onset of symptoms. Three assays suffer from a specificity below 90% (EUROIMMUN IgG and IgA, UNscience, Zhuhai Livzon).

The second week of COVID-19 seems to be the best period for assessing the sensitivity of commercial serological assays. To achieve an early diagnosis of COVID-19 based on antibody detection, a dual challenge must be met: the immunodiagnostic window period must be shortened and an optimal specificity must be conserved.

## I. Introduction

Severe acute respiratory syndrome coronavirus type 2 (SARS-CoV-2) infection was considered pandemic on 11 March 2020. Coronavirus disease 2019 (COVID-19) began affecting France in mid-February 2020, spreading in particular from a large, four-day long evangelical meeting that began on February 17 and gathered 2,500 people in the northeastern city of Mulhouse. The *in vitro* diagnosis of COVID-19 is currently based on the detection of SARS-CoV-2 RNA in respiratory tract specimens (1). Viral RNA can be detected in nasopharyngeal swabs, sputum and bronchoalveolar lavage. However, studies suggest that false-negative test results are relatively frequent, occurring in up to 40% of swab and sputum specimens (2). Confirmation of clinical diagnosis based on molecular tests also can be difficult in mild forms of COVID-19 and when samples are collected long after the onset of the disease (3).

Serology may be a promising way to assess SARS-Cov-2 infection, complementing molecular techniques. Immunological methods can be used to detect the presence of IgM, IgA and IgG directed against SARS-CoV-2 antigenic sites, usually located in the SARS-CoV-2 protein S or protein N (4, 5). Since the beginning of the epidemic, commercial assays - using either laboratory assays or rapid test formats - have been developed rapidly. While serological tests might be a simple and effective screening method, they have shown limitations in the diagnosis of acute infections due to the time required for an adaptive immune response to be acquired. In the very early phase of acute infections, the capacity of serological tests to confirm a diagnosis is hence limited. Subjects with suspected COVID-19 may seek advice and care immediately or several days after the onset of their symptoms. IgM antibodies are produced by short-lived plasma cells during the early phase of the B cell response, providing a first line of adaptive defense against viral infections, whereas the long-term humoral response is based on high affinity IgG. However, the kinetics of the humoral response against SARS-CoV-2 remains incompletely described as it fully relies on the performance of the serological tests used. Serological tests may be useful to confirm SARS-CoV-2 infection when the seroconversion is evidenced, and obviously for epidemiological serological surveys.

Many COVID-19 serological assays are commercialized in Europe, and most are CE-IVD marked. With the exception of high-risk products, whose performance is subject to an external control by a European Notified Body, it is the manufacturers’ own responsibility to ensure that products delivered to European markets meet the essential requirements. The performance of CE-IVD marked assays therefore must be assessed and compared. POC tests dedicated to detecting anti-SARS-CoV-2 IgM and IgG antibodies may also allow access to *in vitro* diagnostic tests outside laboratory facilities. Some authors reported encouraging results using rapid lateral flow assays testing anti-lgM and IgG SARS-CoV-2 antibodies, whereas other studies have reported poor sensitivity in patients with proven COVID-19 (6–8).

In this study, we assessed and compared the performance of six rapid tests and three ELISAs for the diagnosis of COVID-19, and explored seroconversions in subjects with confirmed COVID-19 hospitalized at the Montpellier University Hospital.

## II. Material and Methods

From 18 March 2020, plasma samples were collected from consecutive patients hospitalized in the Montpellier University Hospital with PCR-proven or suspected COVID-19 and included in the “COVIDotheque” cohort (ClinicalTrials.gov Identifier: NCT04347850). Negative PCR-tested patients for SARS-CoV-2 RNA were excluded from the present evaluation of serological tests. The cohort received an institutional ethics committee approval (CPP Ile de France III, n°2020-A00935-34). The demographie and clinical characteristics of the patients are detailed in Table 1. Severity of the Covid-19 infection was defined following current WHO guidelines (6). Controls consisted of samples collected in 2017-2018 in patients care in the department of Infectious Diseases and stored at -80°C until used (DC-2015-2473).

**Table 1:** Patient’s characteristics.

|  | 1 - 6 days* | 7-14 days* | ≥ 15 days* | Controls** |
| --- | --- | --- | --- | --- |
| Number of patients | 9 | 14 | 15 | 20 |
| Age (mean, SD) | 72 (55-90) | 65 (39-86) | 66 (51-83) | 41 (17-72) |
| Sex ratio M/F | 7/2 | 8/6 | 7/8 | 10/10 |
| Severe COVID-19 | 4 | 9 | 13 | - |

\* Days between onset of symptoms and sample collection
\*\* Controls consisted of samples collected in the pre-COVID-19 period (2017-2018)
All serological assays were performed in strict accordance with the manufacturer's instructions.

### ELISAs

The ID.Vet, ID screen^®^ SARS-CoV-2-N IgG indirect (ID.Vet, Montpellier, France), is an assay based on the detection of IgG antibodies directed against the nucleocapsid protein suitable for serum or plasma. A 100 μl volume of 1/20 diluted plasma samples was added to microplate wells and incubated for 45 minutes at 21°C. Wells were washed three times, then HRP-conjugated protein G was added and the mixture was incubated for 30 minutes at 21 °C. Wells then were washed again three times and a chromogen solution was added. Following 20 minutes of incubation at 21 °C, the reaction was stopped and the resultant absorbance was read on a microplate reader at 450 nm. The cut-off value for a positive result was calculated according to the manufacturees instructions: a ratio < 60% is considered negative, ≥ 60% and < 70% borderline, and ≥ 70% positive.

SARS-CoV-2 IgA and IgG (EUROIMMUN, Lubeck, Germany) are assays based on the detection of either IgA or IgG on two separate microplates using a recombinant subunit protein 1 (SI) in serum or plasma. A 100 μl volume of 1:101 diluted plasma samples was added to microplate wells and incubated for 60 minutes at 37 °C. Wells were washed three times, then HRP-conjugated anti-human IgA or IgG were added and the mixture was incubated for 30 minutes at 37 °C. Wells then were washed again three times and a chromogen solution was added. Following 30 minutes of incubation at room temperature, the reaction was stopped and the resultant absorbance was read on a microplate reader at 450 nm with reference at 620 nm. The ratio between the extinction of the sample and calibrator on each plate was calculated. According to the manufacturees instructions, a ratio <0.8 is considered negative, ≥0.8 and <1.1 borderline, and ≥1.1 positive. However, for sensitivity and specificity, 1.1 was used as a more stringent cut-off value for positive results and all values.

### Point-of-care (POC) tests

Six COVID-19 lateral flow assays were evaluated. Five of these detect IgM and IgG SARS-CoV-2 antibodies separately: AccuBioTech Co, Ltd. Accu-Tell^®^ COVID-19 IgG/lgM Rapid Test; (Beijing, China), Zhuhai Livzon Pharmaceutical Group Inc. 2019-nCoV IgM/lgG Antibody Test Kit, (Guangdong, China), Chongqing iSIA BIO-Technology Co., Ltd. 2019-nCoV IgM/lgG Diagnostic Test Kit; (Chongqing, China); UNscience Biotechnology Co., Ltd. COVID-19 IgG/lgM Rapid Test Kit, (Wuhan, China), Acro Biotech, Inc., 2019-nCoV IgM/lgG Rapid Test; (Rancho Cucomonga CA, USA). One only detects IgM: Guangdong Hecin Biotech Co., Ltd. 2019-nCoV IgM Antibody Test Kit, (Guangzhou, China). These POC tests can be performed on whole blood, serum, or plasma, and require 10μl of samples collected by venous puncture or capillary blood collection and a buffer. In the presence of a control signal, any band, even weakly visible, located in the IgM and/or IgG position is considered positive.

#### Statistical analyses

Data were summarized by number and percentage for categorical variables, i.e., positive and negative results. Samples were stratified in three categories according to the delay between the onset of symptoms and sample collection: 0-6 days, 7-14 days, ≥15 days or more. Assay agreement was assessed by computing the percentage of concordant results between tests for each category. Exact 95% confidence intervals were calculated by means of a binomial law accommodating for the small sample size.

## III. Results

Clinical samples collected before the onset of the COVID-19 pandemic were used to check the specificity of the assays (Fig. 1 and Table 2). All results of POC tests were considered as interpretable, although the signal was sometimes weak on the internal control band. IgM and IgG bands were observed in several COVID-19 negative samples using the UNscience POC tests. One out of the 20 COVID-19 negative samples also tested positive using the Zhuhai Livzon POC test as a consequence of a single IgM band. All of the COVID-19 negative samples tested negative using the other four POC tests. False positive results were observed in both the EUROIMMUN IgA and IgG ELISAs. For both of these ELISAs, the results were situated in the “gray zone” in three cases and over the cut-off value in one case. All of the COVID-19 negative samples tested negative using the ID.Vet ELISA.

**Figure 1.**
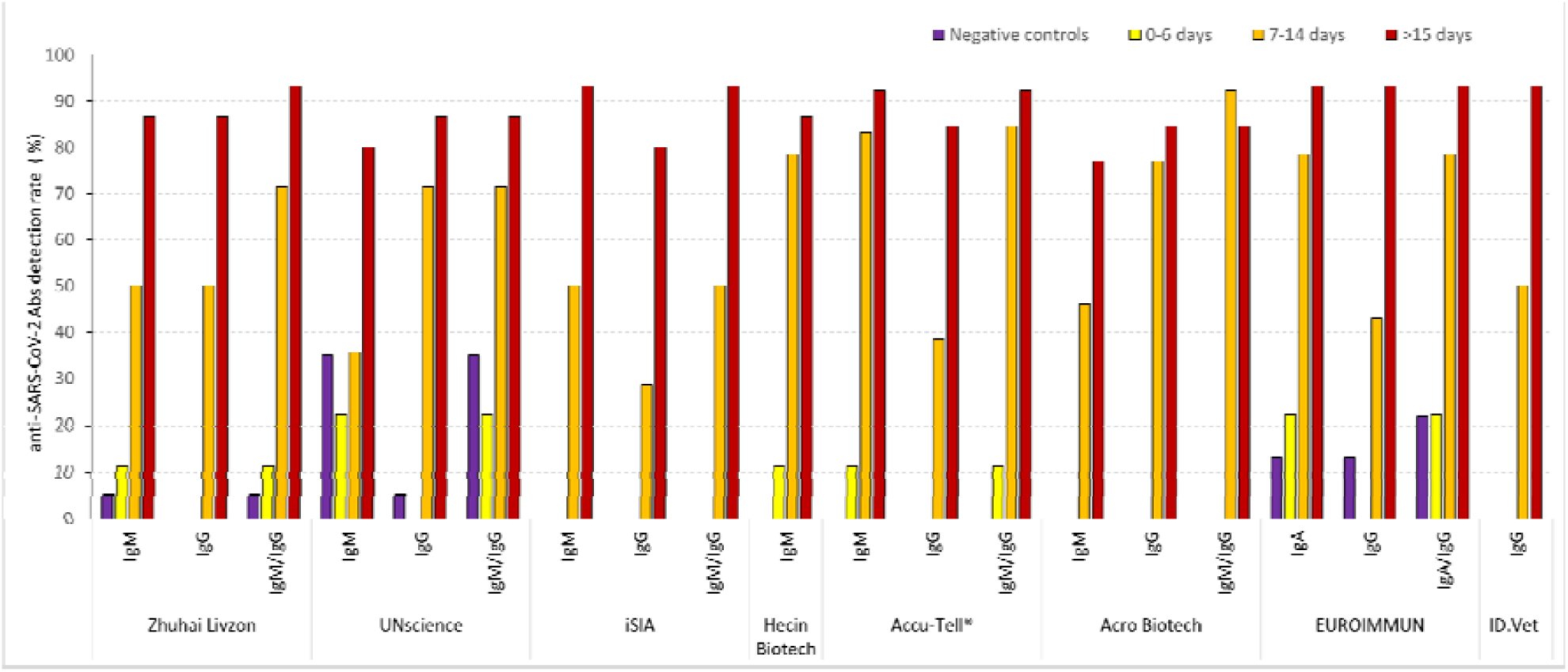
Proportion of samples testing positive for anti-SARS-CoV-2 antibodies. Samples were stratified based on the delay from onset of symptoms. Weak signals, i.e., trace in immunochromatographic test results close to the cut-off value in ELISA, were considered as positive. The proportion of positive tests for each category and for each test and antibody isotype is indicated. For tests combining the detection of two isotypes, the last column indicates positivity for at least one of the two isotypes.

**Table 2.** Performance of the assays observed in the study and provided by the manufacturers.

|  |  | Sensitivity (%)<sup>*</sup> |  | Specificity (%)<sup>*</sup> |  |
| --- | --- | --- | --- | --- | --- |
|  |  | Observed in the study | Reported by the manufacturer | Observed in the study | Reported by the manufacturer |
| **ELISA** | EUROIMMUN (IgA) | 93.3 (80.7-100) | 100 | 80.0 (63.7 - 96.3) | 92.5 |
|  | EUROIMMUN (IgG) | 93.3 (80.7-100) | 80.0 | 85.0 (70.4 - 99.6) | 98.5 |
|  | ID.Vet (IgG) | 93.3 (80.7-100) | 93.3 (78.8-98.2) | 100 | 99.8 (99.3 - 99.9) |
| **Lateral flow assays** | Zhuhai Livzon (IgM) | 80.0 (59.8 -100) | NC | 95.0 (85.4 -100) | NC |
|  | Zhuhai Livzon (IgG) | 86.7 (69.5 - 100) | NC | 100 | NC |
|  | UNscience (IgM) | 80.0 (59.8 - 100) | 98.5 (96.8-99.5) <sup>a</sup> | 65.0 (44.1 - 89.1) | 88.2 (83.1 - 92.2) <sup>a</sup> |
|  | UNscience (IgG) | 86.7 (69.5 - 100) |  | 95.0 (85.4 - 100) |  |
|  | iSIA (IgM) | 93.3 (80.7 - 100) | NC | 100 | NC |
|  | iSIA (IgG) | 80.0 (59.8 - 100) | NC | 100 | NC |
|  | Hecin Biotech (IgM) | 86.7 (69.5 - 100) | 91.3 (87.6-94.2) | 100 | 98.3 (95.8 - 99.6) |
|  | Accu-Tell® (IgM) | 91.7 (76.0 - 100) | 91.8 (83.8-96.6) | 100 | 96.0 (97.7-99.8) |
|  | Accu-Tell® (IgG) | 84.6 (65.0 - 100) | 100 (96.1-100) | 100 | 99.5 (98.1-99.9) |
|  | Acro Biotech (IgM) | 76.9 (54.0 - 99.8) | 85 (62.1-96.8) | 100 | 96.0 (86.3 - 99.5) |
|  | Acro Biotech (IgG) | 84.6 (65.0 - 100) | 100 (86.0-100) | 100 | 98.0 (89.4 - 99.9) |

<sup>\*</sup>in samples collected ≥ 15 days following symptom onset

Samples collected during the first six days of COVID-19 symptoms were rarely reported as positive by any assay, ranging from 0% to 10%, varying with the six assays which all had a 100% specificity. The proportion of samples testing positive for anti-SARS-CoV-2 rose to 50-85% during the second week of COVID-19-related symptoms. IgG bands were more frequently detected or in a similar proportion as IgM bands in POC tests during the second week.

Using the EUROIMMUN assays, anti-SARS-CoV-2 IgA were more frequently detected than IgG during the second week. After at least 15 days following the onset of symptoms, the proportion of anti-SARS-CoV-2 samples exceeded 80% for all of the benchmarked tests. Among the assays that obtained a 100% specificity, two POC tests (iSIA and Accu-Tell^®^) and one ELISA (ID.Vet) obtained a sensitivity greater than 90%. Good overall agreement between assays was recorded during the first week of the disease course as most samples tested negative regardless of the test used, and after the second week of COVID-19 since most of the samples tested positive (Fig. 2).

**Figure 2.**
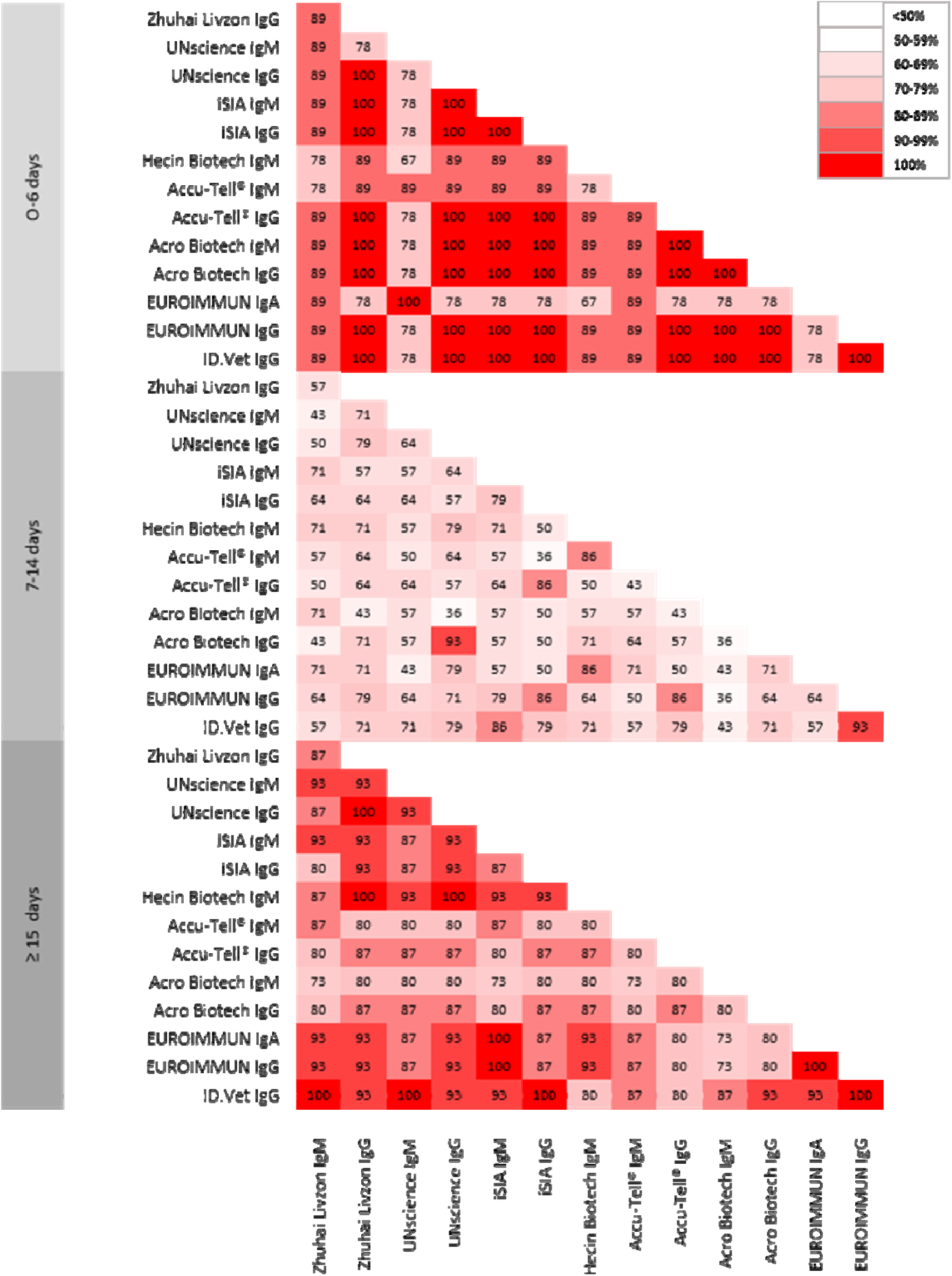
Between-tests agreement rates. The percentage of agreement between the POC and ELISA tests is presented.

Signal-to-cut-off (S/CO) results were analyzed to provide semi-quantitative results using ELISA assays (Fig. 3). We observed a bimodal distribution of S/CO values for anti-SARS-CoV-2 IgG for both the EUROIMMUN and ID.Vet assays. Hence, S/CO remained generally low in samples collected within five days of symptom onset, contrasting most of the time with values from samples collected 10 days after symptom onset that were clearly over the cut-off. Although less obvious, the same trend was observed using the IgA assay. The development of SARS-CoV-2 antibodies was analyzed over time in four hospitalized patients (Fig. 4). In three patients, IgA and IgG were detected by the end of the first week after the onset of symptoms (days 5-7) using the IgG and IgA ELISAs. Development of SARS-CoV-2 antibodies was delayed up to days 12 and 15 for the last patient. Using the Accu-Tell^®^ POC test, seroconversion detected by SARS-CoV-2 antibodies occurred around the same time as the ELISAs. IgG were detectable after IgM for one patients (Fig. 5A), before IgM for one patient (Fig. 5C), and along with IgM for one patients (Fig. 5B&D).

**Figure 3.**
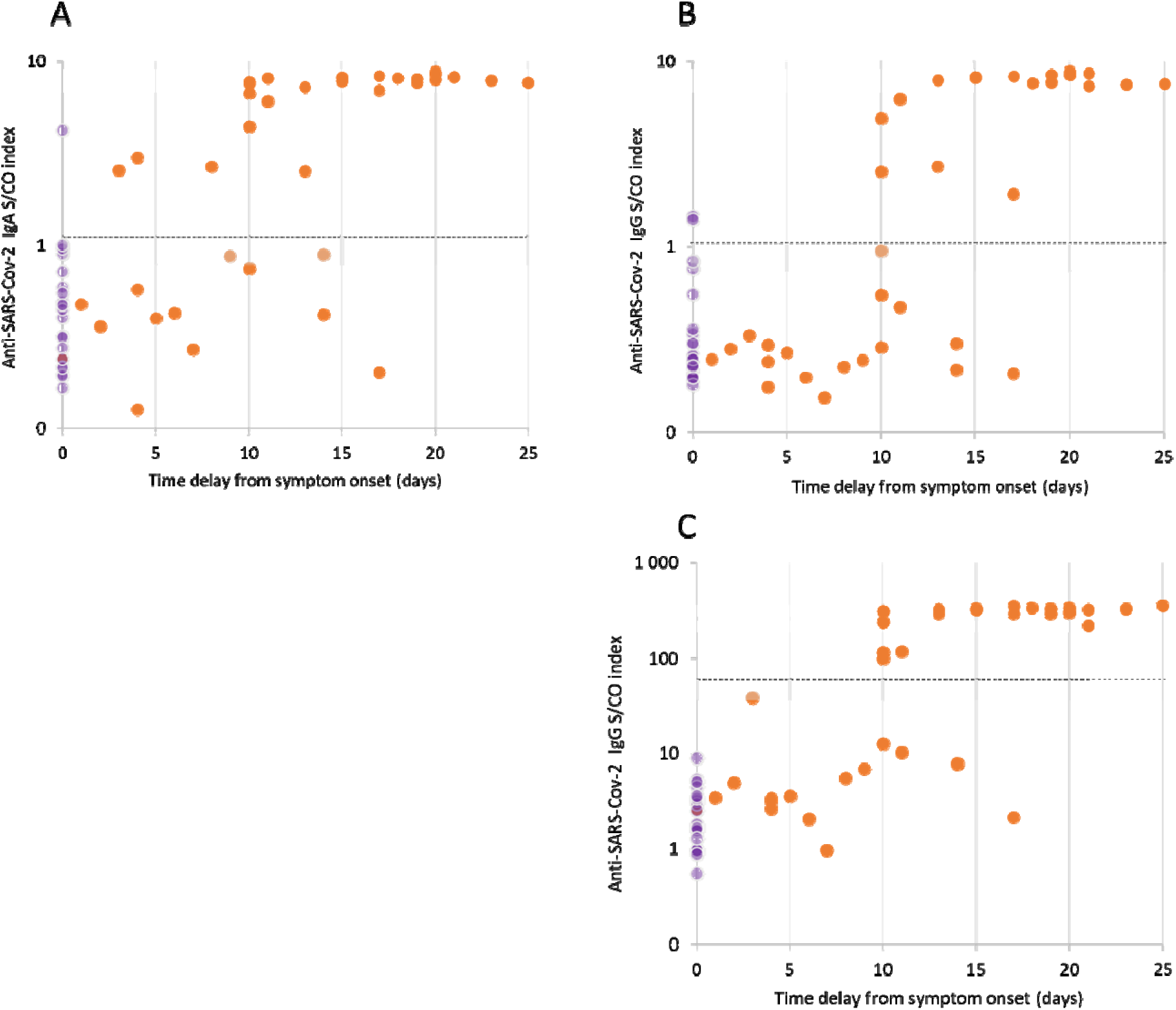
Signal-to-cut-off results according to the time delay from symptom onset. A) EUROIMMUN IgA test. B) EUROIMMUN IgG test. C) ID.Vet IgG test. Samples from patients with proven SARS-CoV-2 infection are indicated by an orange circle, negative controls by a purple circle. The positivity threshold is indicated by the dotted line. Results in the area of uncertainty (gray) were considered positive.

**Figure 4.**
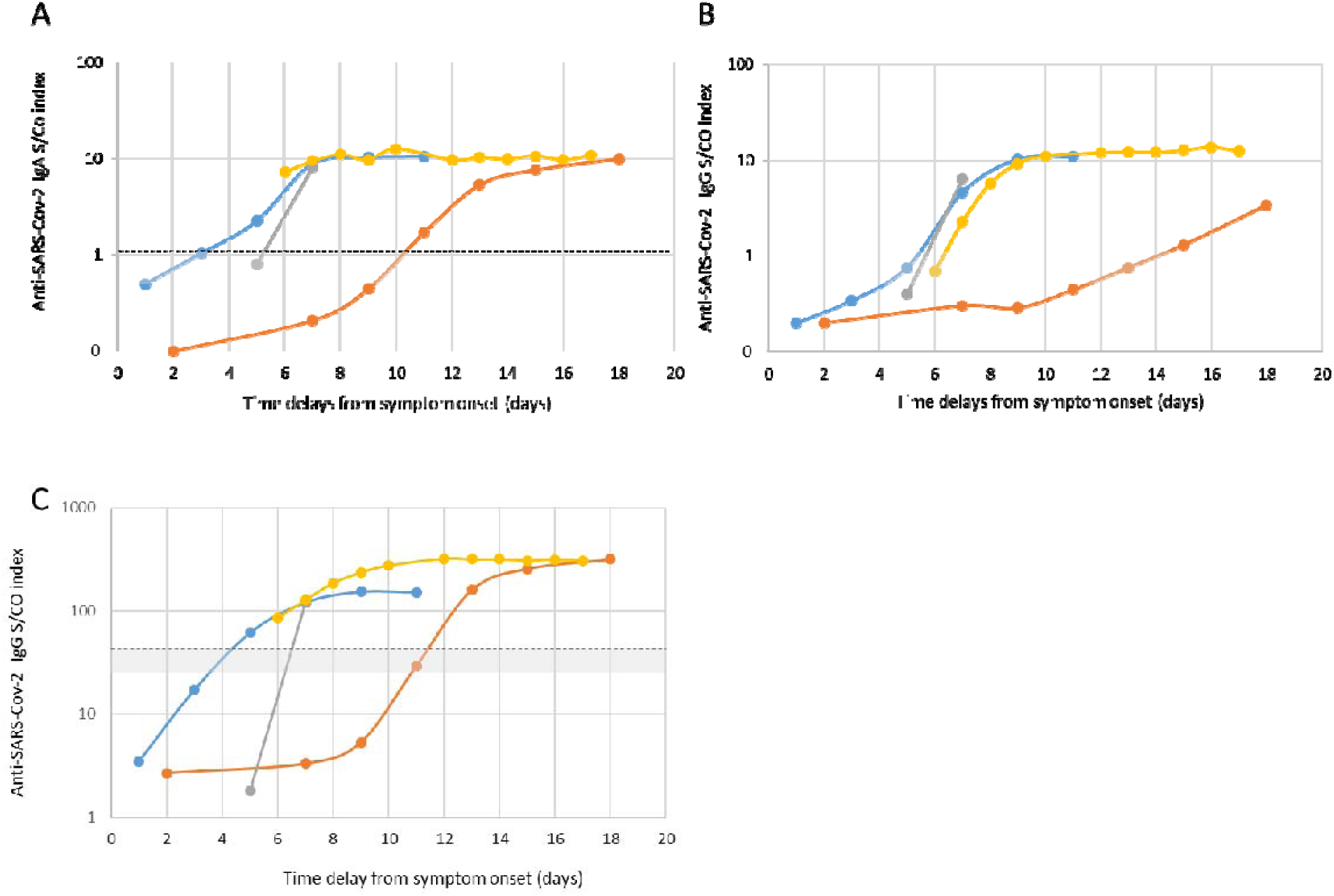
Seroconversions for anti-SARS-CoV-2 antibodies. The results of four patients are presented; in orange patient # 1; in yellow patient # 2, in blue patient # 3, in gray patient # 4. A) EUROIMMUN IgA test. B) EUROIMMUN IgG test. C) ID.Vet IgG test. The positivity threshold is indicated by the dotted line. The area of uncertainty is indicated in gray.

**Figure 5.**
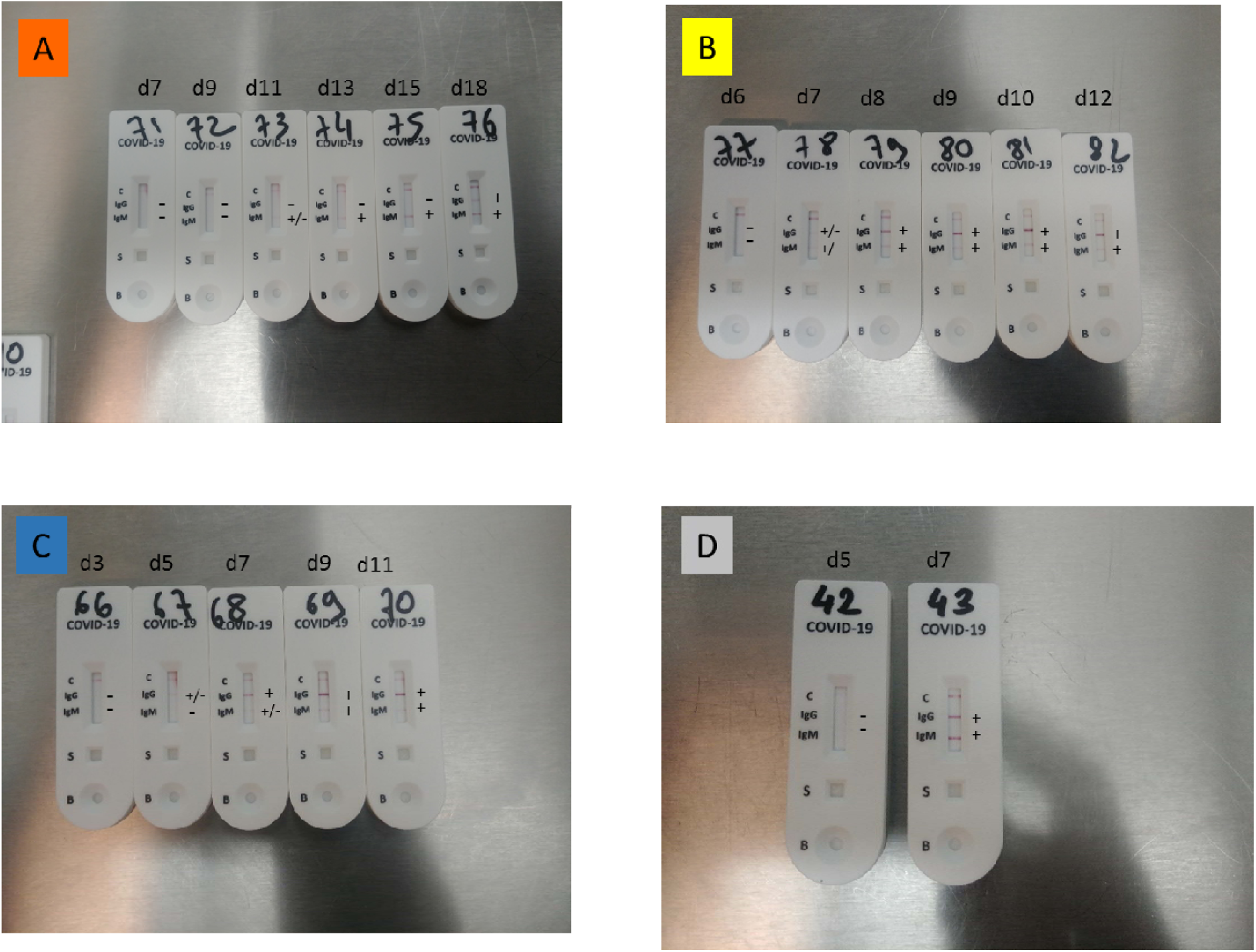
Pattern of anti-SARS-CoV-2 seroconversion using a lateral flow assay. (Accu-Tell^®^). The results of three patients are presented. A) Patient # 1 (orange curve in Figure 3). B) Patient # 2 (yellow curve in Figure 3). C) Patient # 3 (blue curve in Figure 3). D) Patient # 4 (gray curve in Figure 3).

## IV. Discussion

In this study we evaluated the performance of commercial COVID-19 serological assays in POC and ELISA formats. The tests were able to detect anti-SARS-CoV-2 in the majority of samples collected seven days or more after the onset of symptoms. Clinical samples tested during the second week of the disease tested positive with at least two tests. After 15 days, the rate of detection was high (> 80%) but never optimal (<100%) irrespective of the test used. More than 90% of the samples collected more than 15 days after symptom onset tested positive using the iSIA and Accu-Tell^®^ POC tests. Although based only on IgG detection, a similar level of sensitivity was obtained using the ID.Vet ELISA assay. Three assays suffer from an insufficient specificity (EUROIMMUN IgG and IgA, UNscience, Zhuhai Livzon). Repeated testing from the early phase of the symptoms allowed IgG seroconversion to be observed during the second week of COVID-19. These results confirmed previous studies performed in hospitalized patients that used other serological assays (7–10).

Three out of six POC tests had a sensitivity greater than 90% for samples collected at least 15 days after the onset of symptoms. During the second week of COVID-19, IgM were not detected before IgG by means of the POC tests used in this study, suggesting that the IgM bands were not very sensitive. Furthermore, the Hecin Biotech POC test based on detection of IgM alone was not more sensitive on samples collected during the second week of COVID-19 compared to the IgM/lgG POC test. Samples generally tested negative during the first week, suggesting that either this is too early for the anti-SARS-CoV-2 specific response, or the antibody concentration is too tenuous to be detected using POC tests. Considering only the POC tests for which we observed a 100% specificity, IgM bands alone were detected in the few samples that tested positive during the first week of COVID-19. The capacity of POC tests to detect IgM before IgG was disappointing. This observation contrasts with previous studies suggesting that anti-SARS-CoV-2 IgM can be detectable several days before IgG, reducing the window period of serological tests (10, 11). The lateral flow assays tested in this study lack the sensitivity to detect IgM bands, but this may mitigate the risk of non-specific IgM antibody detection due to interference factors such as rheumatoid factor IgM (12). Four of the six POC tests had a specificity of 100%, which will have to be confirmed on a larger number of samples.

We frequently observed a weak signal in the IgM/lgG ELISAs, making it difficult to read the COVID-19 POC tests. By comparison with the POC tests, ELISA offers the advantage of an objective reading. One ELISA manufacturer has based its assays on the detection of antibodies directed against spike protein 1 (SI) antigen (EUROIMMUN), and the second manufacturer on the nucleoprotein (NP) (ID.Vet). The follow up by ELISA of four hospitalized patients confirmed the development of SARS-CoV-2 antibodies at the end of the first week of COVID-19 in three patients, but only after a delay of 12 days in one patient. After 15 days from the onset of symptoms, the three ELISA kits showed a good ability (>90%) to detect SARS-CoV-2 antibodies. As previously reported, anti-SARS-CoV-2 IgA were detected earlier compared to IgG, but the assay had the disadvantage of a poor specificity, which has been reported previously (13). Overall, our results suggest that a second testing of IgA/lgG positive results might be necessary to control the rise of the signal when a low or moderate positive S/CO ratio is obtained when using the EUROIMMUN tests. The prevalence of anti-SARS-CoV-2 was estimated to be as low as 3.1% in the Occitanie region in May 2020 (14). In this context, a very high specificity is required to obtain a good positive predictive value and an accurate estimate of anti-SARS-CoV-2 antibody prevalence in surveys. To control the specificity of the ID.Vet assay, we tested 100 additional samples collected before the occurrence of the SARS-CoV-2 epidemic and obtained a 99.6% specificity (data not shown).

Our study is one of the very first to evaluate the performance of commercial SARS-CoV-2 serologic assays. It suffers two main weaknesses. First, the evaluation is based on a relatively small number of plasma samples. This is due to the fact that only a limited number of samples were available during the early phases of the epidemic in France. As a result, the estimation of sensitivity and specificity values are relatively imprecise. Our preliminary results should be completed by investigations on a larger sample size. Second, we selected samples from hospitalized patients with moderate to severe COVID-19. The intensity of the humoral response to SARS-CoV-2 N or S proteins may be lower in asymptomatic or paucisymptomatic COVID-19 cases. Again, our evaluation should be extended to a larger group of subjects with different clinical presentations including asymptomatic infections.

In conclusion, COVID-19 serological assays in both lateral flow and ELISA formats have a good capacity to detect SARS-CoV-2 antibodies two weeks after the onset of symptoms. The rate of detection is close to zero during the first week. The rate of detection is variable during the second week of COVID-19. Using POC tests, IgM detection did not appear earlier than IgG detection. ELISAs detecting IgG directed against spike protein 1 versus nucleoprotein achieved comparable sensitivities, but with a better specificity for the N protein-based ID.Vet test. The second week of the disease is probably the best period of time to evaluate the sensitivity of the serological assays. Since the most severe symptoms are observed after seven days of evolution, serological assays may be useful in the diagnosis of patients with acute respiratory distress syndrome and a negative PCR assay. In our study, repeated testing confirms that seroconversions occur during the second week of the disease. To achieve an early diagnosis of COVID-19 based on antibody detection, a dual challenge must be met: the immunodiagnostic window period must be shortened and an optimal specificity must be conserved.

## Data Availability

Not available

## Notes

## Acknowledgments

We are grateful to Thomas Landragin for his technical assistance. We thank Grâce Delobel for english language editing and review services.

